# Promoting Vigorous Intermittent Lifestyle Physical Activity (VILPA) in middle-aged adults: An evaluation of the MovSnax mobile app

**DOI:** 10.1101/2024.05.07.24306973

**Authors:** Cecilie Thøgersen-Ntoumani, Anne Grunseit, Andreas Holtermann, Sarah Steiner, Catrine Tudor-Locke, Annemarie Koster, Nathan Johnson, Carol Maher, Matthew Ahmadi, Josephine Y. Chau, Emmanuel Stamatakis

## Abstract

**Background:** Most adults fail to meet the moderate to vigorous physical activity-based recommendations needed to maintain or improve health. Vigorous Intermittent Lifestyle Physical Activity (VILPA) refers to short (1-2 mins) high-intensity activities that are integrated into activities of daily living. VILPA has shown strong potential to improve health and addresses commonly reported barriers to physical activity. However, it is unknown how VILPA can best be promoted among the adult population. This study aimed to evaluate the usability, user engagement, and satisfaction of a mobile application (MovSnax) designed to promote VILPA.

**Methods:** A concurrent mixed methods design was used. It comprised four parts. Part A was a survey with *n*=8 mHealth and physical activity experts who had used the app over 7-10 days. Part B was think- aloud interviews with *n*=5 end-users aged 40-65 years old. Part C was a survey with a new group of 40-65-year-old end-users (*n*=35) who had used the MovSnax app over 7-10 days. Part D was semi- structured interviews with *n*=18 participants who took part in Part C. Directed content analysis was used to analyze the results from Parts A, B, and D, and descriptive statistics were used to analyze findings from Part C.

**Results:** Participants reported positive views on the MovSnax app for promoting VILPA but also identified usability issues such as unclear purpose, difficulties in manual data entry, and limited customization options. Across the different data collections, they consistently emphasized the need for more motivational features, clearer feedback, and gamification elements to enhance engagement. Quantitative assessment showed satisfactory scores on objective measures but lower ratings on subjective aspects, possibly due to unfamiliarity with the VILPA concept and/or technical barriers.

**Conclusions:** The MovSnax app, tested in the present study, is the world’s first digital tool aimed specifically at increasing VILPA. The findings of the present study underscore the need for further app refinement, focusing on clarifying its purpose and instructions, boosting user engagement through personalization and added motivational elements, enhancing accuracy in detecting VILPA bouts, implementing clearer feedback mechanisms, expanding customization choices (such as font size and comparative data), and ensuring transparent and meaningful activity tracking.

## Introduction

The evidence supporting the health and well-being benefits of physical activity (PA) is unequivocal (e.g., (1–3). Current PA guidelines recommend that adults should engage in 150-300 minutes of moderate-to-vigorous PA per week, or 75-150 minutes of vigorous PA, or an equivalent amount to realize these health benefits. However, most adults (especially middle-aged and older) do not achieve these guidelines (4,5). The existence of the well-documented intention-behaviour gap (6) suggests that although adults are aware of the benefits that result in positive intentions to change, these intentions often do not translate into behavioural changes. This apparent paradox can be explained by a myriad of barriers that adults experience in their efforts to increase PA. A systematic review has shown that, among middle-aged and older adults, these barriers include environmental factors (e.g., cost, weather, facilities), individual psychological factors (such as difficulties regulating PA behaviour, doubts about personal capabilities, and lack of motivation), health-related characteristics (e.g., pain, poor health), and opportunity barriers, in particular perceived time constraints (7).

In 2020, the requirement for a minimum bout length for PA was removed from the World Health Organization’s PA guidelines in recognition of emerging evidence that shorter bouts do confer a health benefit (8) and to account for those who report time constraints as a barrier or may not be willing or able to engage in structured exercise. Moreover, accumulation of PA through shorter bouts may be more manageable for individuals facing barriers associated with time and health. The concept of Vigorous Intermittent Lifestyle Physical Activity, referred to as “VILPA” (9), was developed to address these barriers. VILPA refers to brief, vigorous bursts of PA lasting 1- 2 minutes each that are integrated into activities of daily living, such as fast walking to the bus, stair climbing, lifting, or playing actively with children, or walking uphill (9). While other related concepts have been proposed (e.g., exercise snacks; (10)), VILPA distinguishes itself by its focus on integration of specifically vigorous intensity movement into activities of daily living. VILPA is also the only example of vigorous short bout PA that is supported by epidemiological evidence (11,12). Indeed, a recent large epidemiological study focused on non-exercisers identified in the UK Biobank revealed that, compared to individuals performing no VILPA, engaging in three VILPA bouts per day (lasting the equivalent of 1 or 2 minutes each) was linked to a substantially lowered risk of cardiovascular disease (CVD) mortality (by 48–49%) and a marked reduction in all-cause and cancer mortality risk (by 38–40%), and analogous reductions in cancer incidence risk (13) over 7 years. These findings underscore the potential of VILPA as an alternative approach to structured exercise, providing a feasible option for accruing health benefits through brief, vigorously intense, physical activity bouts into daily life.

To date, there are no published studies of interventions designed to promote VILPA. However, a recent qualitative study with physically inactive adults aged 35-76 (*N*=78) provided insight into perceived barriers and enablers of VILPA for this group (14). The findings showed that the barriers and enablers of VILPA differed somewhat to those of other types of PA. The results suggested that barriers to VILPA could be addressed by developing guidelines, addressing safety concerns, and clearly explaining the potential benefits of, and opportunities to perform and accumulate, VILPA. Additionally, VILPA could be promoted by focusing on its time-efficient nature which requires no special equipment or allocated time (e.g., gym sessions) and could be enhanced via the use of prompts and reminders provided at opportune times, and by implementing habit formation strategies (15).

Digital technology is increasingly used as a tool to promote physical activity (16,17).

Smartphone applications (henceforth referred to as mobile apps) have been particularly popular digital tools in the PA field. Given the ubiquitous use of mobile phones, mobile apps have the advantage that they have the potential for wide reach, represent an affordable means of promoting PA, and are a convenient means of engaging many individuals at once. These apps can incorporate a range of behaviour modification techniques designed to change PA behaviour, such as education, individual goal setting, prompts and reminders, self-monitoring, and behavioural feedback. Meta-analytic evidence has provided some support demonstrating the ability of smartphone app-based interventions to promote PA (16–18). One review has shown that factors such as user guidance, health information, statistical progress information, reminders, self-monitoring, goal setting and perceived utility of the app influence the uptake and engagement with health and well-being apps (19).

Mobile apps could be a highly suitable VILPA intervention delivery tool in terms of convenience because people tend to carry their phones with them when performing daily living activities, and because they can provide real-time feedback on behaviour. However, to date, to our knowledge, there are no apps supporting VILPA. Thus, the present study aimed to evaluate the usability, user engagement, and satisfaction of a smartphone app designed to promote VILPA among adults aged 40-65 years old. mHealth solutions like this offer the ability to promote VILPA, helping combat physical inactivity and reducing the risk of chronic diseases, in a scalable and user-centred way.

## Method

### Design

A concurrent mixed methods design, comprising surveys and one-on-one interviews, was used to evaluate the app. The mobile app was evaluated in four parts: Part A) an expert survey (experts were defined as researchers who had published peer-reviewed studies on physical activity and mHealth), Part B) think-aloud interviews with target end-users, Part C) a feedback survey with a new group of target end-users, and Part D) individual interviews with a purposeful selection of participants from c).

### Ethics

Ethics approval for the study (including Parts A-D) was granted by the University of Sydney’s Human Research Ethics Committee (2021/533), prior to the commencement of recruitment of the participants. Participants were informed about the study and had the opportunity to ask questions prior to agreeing to take part. All participants signed written consent forms prior to participating in the study.

### Procedure and Measures

The studies took place from 2022 to 2023.

*Description of the MovSnax mobile app*. A customized app (MovSnax) was developed for the iOS platform with the intention to promote VILPA among middle-aged adults (see Figure 1). As VILPA is an academic term unlikely to be fully understood by the lay public, we presented VILPA to the participants as “vigorous movement snacks” to make it more accessible and tangible for the participants. Decisions about which app features to include were based on reviews of the extant literature on mHealth and physical activity (18,20) and the results of focus groups with members of the target group about barriers and enablers of VILPA (14). The app included automatic detection and daily reporting of VILPA bouts (fast walking and stair climbing), opportunities to set and adjust daily goals, behavioural feedback on daily, weekly, and monthly goal achievement, and dynamic educational and motivational messages which were randomly provided to participants via push notifications in response to periods of inactivity. The app also allowed participants to manually add VILPA bouts, confirm or adjust automatically detected VILPA bouts, enable or disable push notifications and provide feedback on the app. It also collected additional physical activity metrics which were harnessed through the native smartphone app (Apple Health), such as the number of steps accumulated each day. The choice to include educational and motivational messages was based on the results of online surveys with a) researchers with communication expertise, and b) 40– 65-year-old physically inactive adults (results of which are not provided here).

**Figure 1.**
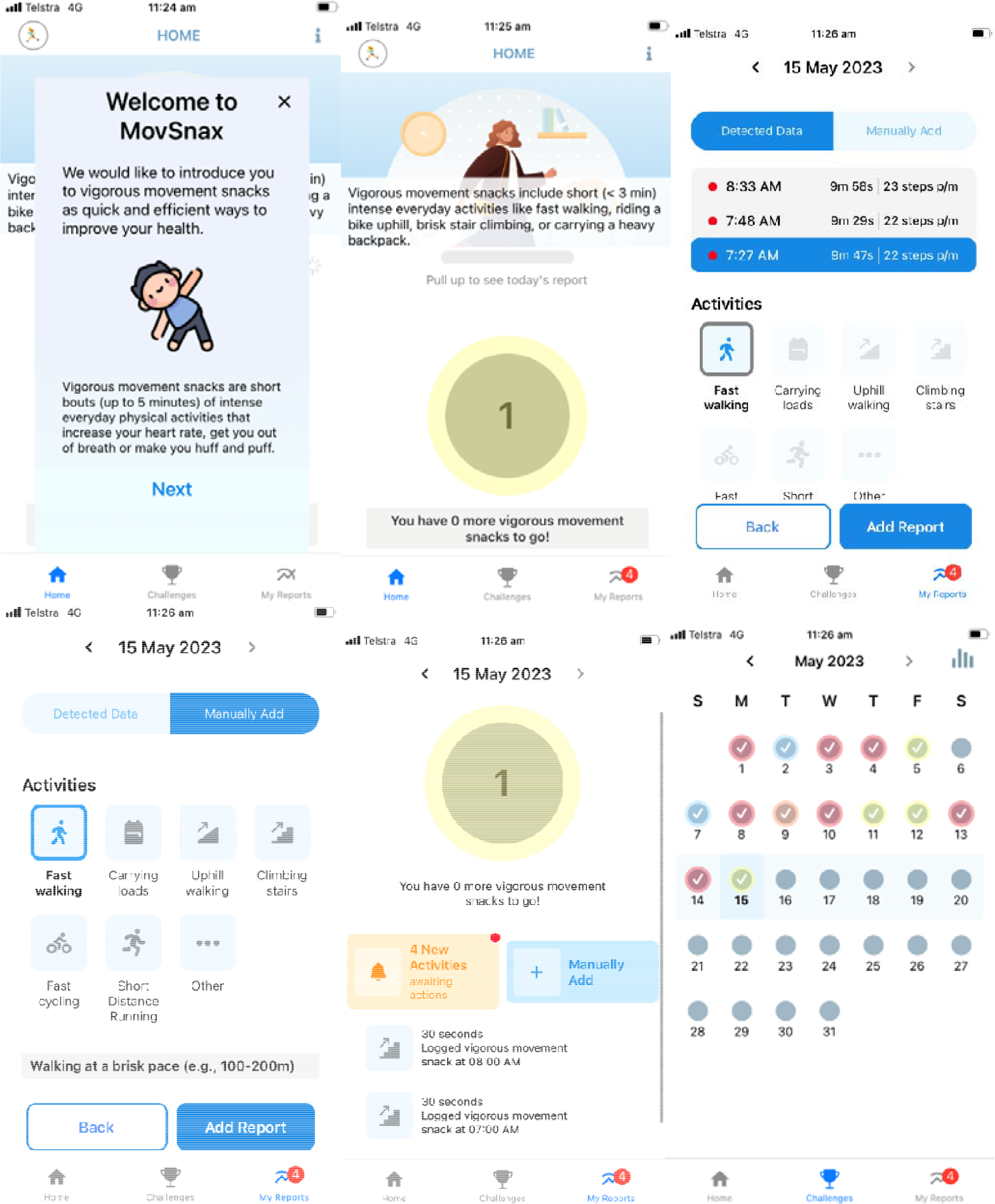
Examples of MovSnax app screenshots

Part A: *Expert survey*. Researchers from the authors’ research networks and contacts with expertise in physical activity and/or mHealth (who had published peer-reviewed studies on physical activity and mHealth) were contacted via email by the research team and asked to volunteer to download the app prototype and use it daily for 7-10 days. Subsequently, they were sent an online survey with open response formats asking them to comment on the app’s functionality, aesthetics, design, perceived usability, benefits, and practicability for the target users (i.e., 40-65-year-old physically inactive adults). They were also asked to comment on what they did and did not like, plus encouraged to reflect on how the app could be improved. The survey is included in Additional File 1.

Part B: *Think-aloud interviews with target end-users*. Adult volunteers aged 40-65 years who were based in Australia were recruited via Facebook ads which targeted people in the age group (40–65- year-old adults) who did not list any sports and recreational activities in their profiles. Upon contacting the research assistant, interested participants were asked to complete a screening questionnaire to assess eligibility criteria that participants: did not take part in a structured exercise program three or more times per week (a similar measure was used in (14)), owned an iPhone; had no mental illness which would preclude participation and were 40 to 65 years of age. Eligible participants who agreed to participate were invited to a one-to-one session with a member of the research team. Four of the interviews were conducted in-person in a meeting room at the University where the study took place, and one interview was conducted online via Zoom with the camera function switched off. The purpose of the session was to get target users’ firsthand impressions of the app. The approach previously used by White and colleagues (21,22) was adopted. The participants were requested to verbally express their thoughts, impressions, and emotions as they explored the app in full for the first time in the presence of the researcher. The interviews were audio-recorded, then transcribed verbatim. No notes were taken during the interviews.

Part C: *Free-living end-user mobile app test with target end-users*. A new group of adults 40-65 years of age were recruited via Facebook ads. The inclusion criteria were identical to those for Part B. Once participants had consented to take part in the study, they downloaded the app, were encouraged to use it daily over a 7–10-day period and were then asked to complete the User Version of the Mobile Application Rating Scale (uMARS) (23) to assess engagement, functionality, aesthetics, information (quantity, quality and credibility), and perspectives on the potential behavioral impact of the app. The uMARS is a 20-item scale which is frequently used to evaluate the quality of apps. The scale provides an overall app quality score (called objective quality) and consists of four sub-scales: engagement (entertainment, customization), functionality (ease of use, navigation), aesthetics (layout, graphics, visual appeal), and information (quality, quantity). A subjective app quality score can also be computed based on questions on the extent to which participants would recommend the app to other people, whether they would pay for the app, and the star-based rating they would give the app. Finally, perceived impact was computed based on answers relating to questions on the extent to which the app increased awareness, knowledge, changed attitudes and intentions to change, induced help seeking, and the likelihood of the app promoting behaviour change. Each item was rated on a scale from 1 to 5, with 1 being the least desirable (e.g., not at all/strongly disagree) and 5 being the most desirable (e.g., definitely/strongly agree). A score of 3 has been described as satisfactory (24,25). Evidence of the scale’s internal and test-retest reliability has been provided in previous studies (23).

Part D: *Interviews with target end-users from Part C*. Volunteer participants were purposefully selected to explore their views from the survey (Part C) in more detail. Purposeful sampling was used whereby participants representing different demographics, and who had more, and less favourable views of the app were selected. All interviews were conducted via Zoom with the camera setting switched off. Questions pertained to their experiences of using the app, what they liked and disliked about the app, which functions of the app they used, which features were least versus most useful, and how they thought the app could be improved. The interview schedule is presented in Additional File 2. The interviews were recorded via the audio function in Zoom, then transcribed verbatim. No notes were taken during the interviews.

At the end of Part D, the app was revised to incorporate the results from Parts A-D.

### Data analysis

Directed content analysis (26) was used to analyse the open-ended survey responses from experts (Part A), the think-aloud interviews (Part B), and the final interviews (Part D). Codes, which are short descriptive labels, were created to organize and understand the data. The first author read and re-read the data to become familiarized with the data. She then inductively developed the codes and sub-codes and provided example quotes for each. The second and penultimate authors read through a selection of the Part B and D interviews and critically appraised the codes and sub-codes developed by the first author. This resulted in the revision of some of the codes and sub-codes. Part C data were analysed using descriptive statistics.

## Results

*Part A.* Twenty experts signed the consent form. Of those, eight researchers (*N*=3 females, *N*=4 males, *N*=1 unknown) downloaded the app, used it in their daily lives for 7-10 days, and then completed the online expert survey. The participants engaged in *M* (SD) = 9 (11) (range 4-41) vigorous movements snacks per day during the testing phase. The survey results from this stage are reported in detail in Additional File 3. We created six codes: Purpose, design, ease of use, user engagement, tracking and reporting, and education and knowledge. Overall, the participants reported finding the app visually appealing and relatively easy to use. However, there were concerns about the lack of clarity about the purpose of the app and a lack of instructions about how to use the app effectively. User engagement was a prominent theme with several suggestions provided for how the app could be improved to increase engagement. These included allowing for more personalization, adding motivational features (such as gamification) beyond monitoring, and adding push notifications to remind and otherwise engage the users with the app over the longer term.

The automatic detection of vigorous movement snacks was generally viewed by the respondents as a strength of the app, although several participants stated that the reporting of the VILPA bouts was often inaccurate. This could, however, at least partly reflect that the participants might have done VILPA without realising it. Two participants mentioned that accurate syncing between the app and other fitness trackers was needed. One participant also noted difficulties accessing basic data such as step counts. Another participant noted that they found it frustrating having to manually confirm automatically detected data. The need to improve the accuracy of the automatically detected VILPA bouts without having to manually confirm these appeared important to several participants. Finally, while educational messages were included in the app, some participants mentioned that the app should include short educational videos and provide more information about the importance of doing daily VILPA. Overall, the analysis of the data from Part A revealed that the app had several positive features that the end-user might find useful. However, the results also demonstrated that improvements could be made to enhance user engagement, motivation, and overall user experience.

*Part B.* Five target end-users (3 females, and 2 males) took part in the think-aloud interviews. All participants completed all tasks on the walkthrough checklist. Additional File 4 presents the coding framework and the specific findings. Seven inductive codes were developed: Onboarding process, user experience, user interface design, education, data detection, recognition and rewards, and customisation. The findings highlighted several issues and suggestions regarding the password setup process and user interface of the app. Users expressed frustration with the current method of password validation, advocating for immediate notification of criteria non-compliance to avoid repetitive data entry. Additionally, there was a desire for clearer guidance on password requirements and parameters. Furthermore, users offered feedback on various aspects of the app’s functionality, including customization options, iconography, and colour scheme, suggesting improvements for a more user-friendly experience. Other feedback centred on the need for clarification on terminology as well as features related to goal setting, data tracking, user profiles, and being acknowledged for longer-bout activity that did not meet the VILPA criteria. Overall, the findings underscored the importance of refining the app’s interface and functionality to enhance user satisfaction and engagement.

*Part C.* Of 54 end-users who provided written informed consent to participate, *N*=35 target end-users (*N*=28 females, 7 males; age range = 40-65; *M* (SD) age = 54.09 (7.57)) downloaded the app, used it daily over a 7–10-day period and then completed the uMARS (23). The socio-demographic and health characteristics of the participants are presented in Additional File 5. On average, the participants completed 4 (*SD*=4; range: 1-16) VILPA bouts per day during the testing period. The results on perceptions of app quality as rated by the uMARS are presented in Table 1. The objective quality score was the highest overall (an above-average rating), while the subjective rating was the lowest. In terms of the objective sub-scale, the participants rated the app highest for aesthetics and information, and the lowest rated subscale was for engagement. Although the subjective quality ratings were fairly low, perceived impact, awareness and behaviour change were rated above the score 3 which have been previously defined as satisfactory (24,27).

**Table 1.** Part C app quality rated using the uMARS (*n*=35)

| **Item** | ***M (SD)*** |
| --- | --- |
| **Objective quality subscale (range: 1-5)** | 3.32 (.77) |
| Engagement | 2.81 (.88) |
| Functionality | 3.26 (1.08) |
| Aesthetics | 3.47 (.90) |
| Information | 3.43 (.85) |
| **Subjective quality total score (range: 1-5)** | 2.43 (1.23) |
| What is your overall (star) rating of the app | 2.51 (1.27) |
| Would you recommend this app to people who might benefit from it? | 2.37 (1.19) |
| How many times do you think you would use this app in the next 12 month? | 2.91 (1.80) |
| Would you pay for this app? | 1.91 (1.15) |
| **Perceived impact (range: 1-5)** | 2.98 (1.17) |
| Awareness | 3.14 (1.22) |
| Knowledge | 2.97 (1.25) |
| Attitudes | 3.06 (1.33) |
| Intention to change | 3.03 (1.34) |
| Help seeking | 2.57 (1.17) |
| Behavior change | 3.09 (1.25) |

*Part D.* Eighteen participants from Part C signed the consent form, and *N*=15 (*n*=11 females, *n*=4 males; age range=40-63) agreed to take part in the interviews. Additional Table 6 presents the coding framework and specific findings. We constructed 5 codes: App concept, user experience, customization, user engagement, and transparent and meaningful tracking.

*App concept*. Participants generally had positive views on the app’s concept, recognizing its potential value despite acknowledging the need for improvements. They appreciated its focus on various aspects of movement beyond step count, such steps per minute, and its ability to acknowledge small efforts in physical activity. Some participants expressed enthusiasm for the idea and willingness to use it once operational issues were resolved, and others believed it was a good concept with potential once tweaks were made. Overall, participants were positive about the app’s concept while recognizing the importance of refinement for its effectiveness.

*User experience*. Participants had mixed views on the usability and effectiveness of the app’s interface. Some praised its simplicity, clean design, and ease of navigation, finding it pleasant, user-friendly, and motivating. They appreciated features like clear notifications and the ability to view data intuitively. However, others criticized the lack of explanatory information, difficulty in understanding functionalities like manual data entry, and overwhelming amount of data without clear direction or value. They expressed frustration with the interface’s limitations, including the inability to customize or expand features, leading to a perception of minimal gain for significant effort. Overall, while some found the interface intuitive and motivating, others struggled with its usability and felt it lacked essential explanations and functionalities.

*Customization*. Participants expressed dissatisfaction with the lack of customization options in the app, particularly regarding font size, which posed challenges for users with specific accessibility needs. They also highlighted the absence of comparative data with peers or benchmarks based on age or other demographics, which they believed would enhance motivation. Additionally, they emphasized the importance of considering age-related factors such as text size for readability, suggesting that tailored features could improve user experience and engagement, particularly for older individuals. Overall, participants desired more flexibility in customization and comparative data to personalize their experience and enhance motivation to use the app.

*User engagement*. Participants noted that while the app kept them entertained and motivated to achieve their goals, they felt that it lacked some features which conferred rewards. Some credited the app with motivating them to do VILPA bouts they might not have otherwise done. However, they also expressed a desire for gamification, for example, more tangible rewards, such as a system of reward points to incentivize participation, which would motivate them for sustained use of the app over the longer-term. Additionally, participants highlighted the importance of clear feedback mechanisms within the app, such as congratulatory messages or visual cues like fireworks upon goal completion, to enhance their sense of achievement and motivation. They suggested incorporating more interactive features like prompts and notifications to encourage consistent engagement and goal attainment. Gamification was mentioned as a potential mobile app feature that could keep participants engaged.

*Transparent and meaningful tracking*. The participants expressed mixed feelings about the amount of data collected. While some appreciated the app’s sensitivity in picking up various movements and generating a significant amount of data, others found it overwhelming and lacking in meaningfulness, particularly when faced with a high volume of recordings. Some noted instances in which certain activities were not accurately counted, which led to frustration and confusion regarding the app’s tracking mechanisms. Despite these concerns by some participants, others reported no issues with the tracking and believed it accurately recorded their activities. Overall, the participants highlighted the importance of striking a balance between sensitivity and meaningfulness in data collection, as well as ensuring accurate tracking to enhance the user experience and satisfaction with the app.

## Discussion

The purpose of the present study was to evaluate a customized mobile app, MovSnax, designed to promote VILPA (referred to as vigorous movement snacks among participants) among middle-aged adults 40-65 years of age. The evaluation of the MovSnax mobile app, designed to promote VILPA among middle-aged adults, has provided insights into its potential effectiveness and areas for improvement. Our findings reveal an interplay between the app’s perceived strengths and the opportunities for enhancement identified by users and experts. Participants acknowledged the app’s aesthetic appeal and informative content and appreciate its innovative approach to promoting physical activity through VILPA. However, the feedback highlighted several areas needing refinement, such as enhancing user engagement, improving the accuracy of activity tracking, and expanding customization options. Despite the generally positive perceptions of the app’s concept and functionality, both the end-users and experts emphasized the need for clearer instructions, more interactive features, and a deeper integration of motivational elements to foster sustained app usage and, ultimately, a more active lifestyle.

Overall, the data suggested that the participants viewed the concept of the app positively, despite some identified issues with its usability and functionality. They appreciated the app’s focus on integrating VILPA into daily activities, aligning with the growing interest in this approach as an alternative to structured exercise, especially for those with time constraints and health concerns.

However, participants also noted areas for improvement in the app’s user interface, particularly regarding clarity of purpose and difficulties in manual data entry. They expressed a desire for more customization options and comparative data with peers. Additionally, participants stressed the importance of incorporating motivational features and rewards to enhance engagement and adherence to VILPA. While the app was praised for motivating short vigorous intensity activities, some participants wanted more tangible rewards (e.g., reward points and leaderboards) and clearer feedback mechanisms. These suggestions align with the results of a systematic review on the factors that facilitate the uptake and engagement with health and well-being smartphone apps (19).

Gamification was also mentioned by both the end-users and the experts as a possible feature that could help engage the users. Indeed, meta-analyses have highlighted gamification, which uses game design elements (e.g., points and leaderboards), as a promising feature in the promotion of physical activity (28,29). The study also revealed mixed feelings among the participants about the amount of data collected by the app, with some finding it overwhelming and lacking in meaningfulness. This underscores the importance of striking a balance between sensitivity and meaningfulness to users when harvesting data.

Regarding the quantitative data, in Part C, the MovSnax app scored over 3 (on a 5-point scale) on the overall objective quality dimension of the uMARS, and its sub-components functionality, aesthetics, and information. The app also received scores of over 3 on perceived impact relating to raising awareness (of VILPA), encouraging behaviour change and changing attitudes plus increasing intentions to increase VILPA. As scores of 3 and above on the uMARS have previously been described as satisfactory (24,27), our results suggest that the app was satisfactory for some criteria (overall objective quality dimension, functionality, aesthetics, and information, and impact) and less than satisfactory (<3) for others (subjective quality, including, e.g., star-based ratings of the app, and the extent to which participants were willing to pay for the app). The relatively low scores on subjective app quality overall could be due to two reasons. First, MovSnax tested a new physical activity concept that the participants were unfamiliar with. The qualitative data suggested that many participants struggled to understand the purpose of the app and that it lacked guidance and clarity, including information about what qualified as a VILPA bout. Second, the qualitative data revealed a diverse number of barriers, many of which were technical, which participants experienced in their use of the app. The lowest rated item was on their willingness to pay for the app (*M*=1.91). This could be partly explained by the large number of freely available fitness and exercise apps, creating expectations of mobile apps being provided at no cost.

To our knowledge, MovSnack is the first mobile app anywhere in the world, aimed at support VILPA. Strengths of the study include its grounding in a solid evidence base of formative work with the target population, ensuring it addressed their stated needs and preferences. Furthermore, we used a mixed-methods design, which allowed for a comprehensive evaluation of the mobile app and triangulation of data. Another strength was that we included both end-users and experts in the multi-stage evaluation. Combining these perspectives allows for an identification of a wider range of strengths and weaknesses, leading to more informed decisions for app refinement and enhancement. Finally, we used a standardized tool to assess app quality and user perceptions, which facilitates comparisons with existing research findings and promotes transparency in reporting the results of app evaluations. However, some limitations associated with the app and the study design also need to be considered in the interpretation of the study’s findings. First, due to time and other practical constraints, it was not possible to test the app iteratively. Further, the app was developed for iOS platforms, possibly limiting the generalizability of the findings. However, iOS is the leading mobile operating system in Australia with 60% of the market in 2023 (30). Finally, app usage data (how long and how often the participants interacted with different features of the app) could not be retrieved due to technical problems.

The study’s findings hold implications for the broader field of mobile health, particularly in the domain of promoting physical activity amongst middle-aged adults. The insights gained from the MovSnax app evaluation illuminate the critical balanced required between technological innovation, user engagement, and practical functionality in app design. It underscores the need to embed motivation psychology within the app’s framework, suggesting that future mHealth tools should integrate personalised, adaptive features, that resonate with users’ lifestyles and preferences, thereby promoting sustained engagement. Future research is needed to refine the app features and functionality incorporating user feedback. Evaluation using an experimental, controlled trial design will then be warranted, to determine the app’s impact on VILPA behaviour change, and sustainment of such change. More broadly, integrating advanced data analytics and machine learning could offer personalised insights to users, potentially revolutionising the way VILPA interventions are delivered and experienced.

## Conclusion

The mobile app evaluated in the present study represents the world’s first (digital) tool designed to promote VILPA in middle-aged adults. The results of a mixed methods evaluation with mHealth and physical activity experts and target users suggest some future potential of the MovSnax app. However, our findings also suggest that the app could be refined by addressing concerns about clarity of purpose and instructions, improving user engagement through personalization and the provision of additional motivational features, enhancing the accuracy of automatically detected VILPA bouts, providing clearer feedback mechanisms, offering more customization options (e.g., font size and comparative data), and ensuring transparent and meaningful tracking of activities. The findings from the present study pinpoint specific areas to enhance the MovSnax app. Future research should utilize these insights to develop and assess an improved app version, followed by feasibility studies and larger trials to evaluate long-term user engagement, behaviour change outcomes, and user experiences through qualitative research. Once refined, the MovSnax app may be used as one component of complex interventions designed to promote VILPA in middle-aged adults, who are unwilling or unable to take part in structured exercise.

## Supporting information

Declarations

## Data Availability

The datasets used and analysed during the present study are available from the corresponding author upon reasonable request.

