## Supplementary material for "Promoting Vigorous Intermittent Lifestyle Physical Activity (VILPA) in middle-aged adults: An evaluation of the MovSnax mobile app": Table S1

Table S1. Part A survey questions

| |
|---|
| Which of the following functions did you explore (tick options)? Weekly progress, Adjusting daily VILPA goals, Notifications, VILPA self-reporting, Steps tracking, Others |
| What did you think about the functionality of the app? |
| What did you think about the aesthetics or design of the app? |
| How useful do you think the app will be in promoting VILPA in 40-65-year-old adults who are physically inactive?<br>In your response, please comment on specific functionalities of the app. |
| How relevant do you think the app will be in promoting VILPA in 40-65-year-old adults who are physically inactive?<br>In your response, please comment on specific functionalities of the app. |
| How beneficial do you think the app will be in promoting VILPA in 40-65-year-old adults who are physically inactive?<br>In your response, please comment on specific functionalities of the app. |
| Please describe your overall experience of using the app over the past 7 days – Aesthetics |
| Please describe your overall experience of using the app over the past 7 days – Practicality |
| Please describe your overall experience of using the app over the past 7 days – Functionality |
| Please describe your overall experience of using the app over the past 7 days – Usefulness |
| What did you like the most about the app? |
| What did you not like about the app? |
| How do you think the app could be improved? |
| Are there any other comments you would like to make about the app? |
