## Supplementary material for "Promoting Vigorous Intermittent Lifestyle Physical Activity (VILPA) in middle-aged adults: An evaluation of the MovSnax mobile app": Table S2

Additional File 2. Table S2. Part D interview schedule

| |
|---|
| First, can you tell me a bit about your experiences of using mobile apps in general? |
| And what about apps designed to increase physical activity |
| Can you tell me about your experiences of using the VILPA app? |
| (following up on their responses to the uMARS survey): Your score on X [e.g., functionality] was fairly low– can you elaborate on your reason for this score? |
| In contrast, in the survey, you scored Y [e.g., engagement] quite high – what was your reason for that? |
| What, if anything, did you like about the app? |
| What, if anything, didn't you like about the app? |
| Did you choose to explore some functions and not others? If so, which and why? |
| If *the participant indicate they didn't use one or more functions much: what were some reasons or barriers to using them?* |
| Which functions you did not explore at all? If so, which and why? |
| How helpful do you think the VILPA app would be to increase your levels of VILPA? In what ways? How? |
| Which parts of the app would be more/less helpful? |
| How do you think the app could be improved? |
| Would you be recommending the app to others? Why, why not? |
