## Supplementary material for "Promoting Vigorous Intermittent Lifestyle Physical Activity (VILPA) in middle-aged adults: An evaluation of the MovSnax mobile app": Table S3

Table S3. Part A coding framework and findings

| Codes | Subcodes | Quotes |
|---|---|---|
| Purpose | | I did not know what a "vigorous snack" was (P2) |
| | | I found it difficult to understand what I was meant to do...I was confused on the purpose of the app and how it would keep people engaged over time (P3) |
| | | I didn’t really understand how the app worked (P8) |
| Design | App appearance Aesthetics | It looks good, it’s very clean and the pictures/icons are appropriate (P1) |
| | | Colours were pleasing (P3) |
| | | I think the design is clean, simple, yet highlights key aspects of the app such as the daily target (P7) |
| | | Nice and clean… Little graphics were cool! (P8) |
| Ease of use | | |
| | User interface | I couldn't find the weekly progress button, and I wasn't aware of it until the start of this survey when it was listed. (P1) |
| | | I also struggled to find where my total step count per day was… Generally, total steps in an easy to find location would be more appropriate. (P2) |
| | | I was not sure…what I was supposed to do (P3) |
| | | The app was fairly easy to navigate, between the tabs, and reporting activity straightforward (P4) |
| | Challenges tab | I found it difficult to navigate to the different challenges (P2) |

| | | |
|---|---|---|
| | | The Challenges section was just identifying a calendar with your daily notice of whether you met your goal or not (P3)<br><br>The challenge tab is the key to this app's success. The age group 40-65 are still motivated with easy uptake of tech and apps - so, incorporating a rewards system may help in incentivising the participants (P6) |
| | Push notifications | …could be very useful with more push notifications to home screen (P2) |
| User engagement | Measurement tool Not a motivational, tool | I am not entirely sure the app came across as a promotional tool, but rather a logging tool… Interesting to self-monitor or log behaviour, not convinced it is motivating (P4) |
| | Gamification<br><br>Individualisation | The motivation to engage in VILPA, to me, may need to come from other sources, or have more gamification built into the app so that people are engaged with the tool and behavior as a means of scoring points, earning rewards or badges, social challenges or leaderboards etc. (P4) |
| | Motivational messages | I regularly completed my self-set goal of 4 vigorous activity snacks and it would have been sensible for the app to acknowledge my achievement and suggest I make the goal more challenging? (P7)<br><br>I wonder if during set-up there needs to be more individualisation of the user to the app to help recognise what counts as a movement snack for them (P7)<br><br>Although self-monitoring can help promote physical activity, there is opportunity with the app to integrate other features beyond self-monitoring such as optional motivational messages, reassurance if a goal is not achieved, support confidence for physical activity, remind users why engaging in movement snacks is valuable and so forth (P7)<br><br>Adding push notification would have helped to remind me to check in (P8) |

| | | |
|---|---|---|
| Tracking and reporting | Detection of activity<br>Syncing<br>Data accessibility<br>Accuracy | The syncing with my activity tracker was not accurate in terms of "activity snacks" (P3)<br><br>Automatic detection of activity - very cool! (P4)<br>(need) more easily accessible data and an end-of-day or weekly report (P2)<br><br>Having to confirm detected data for it to contribute to the report was an annoyance. Often when confirming detected data I was told you haven't selected an activity type, when you think you have, also does not help - you want the app to be automatic and seamless, not feel like hard work (P7) |
| Education and knowledge | Content<br>Education | …an education section with short videos explaining some of the key concepts. For people who don't really understand health information it would be useful to include more background information to support their understanding and the relevance of the app (P1)<br><br>I think the App will need to be accompanied by information on why VILPA is important, and the utility of goal setting and self-monitoring, to increase buy in (P4)<br><br>I wonder if during set-up there needs to be more individualisation of the user to the app to help recognise what counts as a movement snack for them (P7) |

*Note*. P refers to participant ID
