## Supplementary material for "Promoting Vigorous Intermittent Lifestyle Physical Activity (VILPA) in middle-aged adults: An evaluation of the MovSnax mobile app": Table S4

Table S4. Part B coding framework and findings

| Codes | Subcodes | Quotes |
| --- | --- | --- |
| Onboarding process | Password sludge | Yeah, it's just a bit annoying because it should tell you straight away if your password is not good. Instead of having you enter everything and then letting you know so you have to go back and then do everything again Notify user immediately if password does not meet criteria instead of after entering personal information (P2)<br><br>It would be nice if, like if I didn't put in the right password at the beginning, then it asked me to do it right. Yeah, rather than having to fill this out again and then (P3)<br><br>So that's the problem, though it's hard to get set a password, I would have thought this isn't doesn't need such a level of privacy that you need such a complex password. (P5)<br><br>And it doesn't let you see what the password is… (P5) |
| | Guidance and instruction | There wasn't anything confirming like what I'm about to go into so (P3)<br><br>And then you set your own parameters, but it doesn't guide you what's right for your age or your level of fitness, you know (P5) |
| | Remind users of purpose | …thought that it would be useful to have a place in the app where we could remind people of what we mean by movement snacks (P4) |
| | Challenges setting up account | So sometimes when I type in here… It's hard to get back out. (P3)<br><br>it's hard to find that app, yeah… So yes, this this is a problem straight away that I can't find it, so I'm not gonna use it, right? (P5)<br><br>So in the settings you can choose push notifications…. I'm not sure how to find out what that is (P5) |

| | | |
|---|---|---|
| User experience | Avatar, icon and emoji preferences | I like the cute little avatars (P1)<br><br>And this sort of avatars…I don't think that's silly. That they're right. It's just more options would be nice, that's all (P2)<br><br>Cute little character here (P3)<br><br>…also with the icons. The icons are a little bit silly (P4)<br><br>…if I wanna use it I can put an emoji or faces things like that because it resembles a person, especially the ones that you have that example there …the guy running or… things like that more options, but it's about more options I guess. Photos, emojis (P2) |
| | App name perception | Initially, when I heard the word movement snacks was like it's a funny name because like snacks is like food (P4)<br><br>Is that (move snacks) a term that you've made up or is there an existing term? Why did you make up something new? (P5) |
| | Adjusting goals | I wonder if you know, if you can get the app to recognise like they've had three days where they've done 5 and then it prompts…and it says, wow, you're doing amazing. You've had three days above your target, your current target. Why don't you do you want to set a new baseline? (P3)<br><br>And then like having that little explanation which is good from the start and if it gets maybe like it's a good start, they'll go of at least five is great. You can adjust this goal now or in your profile settings (P4) |
| | Privacy concerns and personalization | Put that date of birth, but maybe that's too identifiable for people to give away that instead of, you know, putting your age in that, you know, recalculates every time, give you happy birthday, you know, things like that (P2) |

| | | |
|---|---|---|
| User interface design | Colour scheme | I like the colours, so nice and common and simple to follow (P1)<br><br>Like if you could get really really cool bumpy like colours and…Because it just now looks like alright, it's just very simple and easy, but I think if in the future you would roll it out more widely I would use like some funky (colours) (P4) |
| | Visual feedback | Maybe with that calendar, with the dates where you've got all the other days of the month, maybe make the circles a bit darker. To our a bit more contrast in colour, perhaps, instead of like the blue on the blue, maybe a little bit, maybe darker blue wall, something something a little bit more standing out (P1)<br><br>OK, I would be going through and picking all the ones you want to add and then those ones get ghosted out or it turns green, the dot turns green so you know you've added them (P5) |
| Education | Raising awareness | At a certain pace or a certain time frame, it would be interesting to see like which ones qualified and which ones didn't (P3) |
| | Exemplify vigorous movement snacks | More just to fulfil my challenge, people might want to do something like that, like little examples of exercises that I can just go and do it right now in my room and I can just, you know, fulfil the goals (P2) |
| Data detection | Clarify detection of data | I think it's a really good thing to have there, but it it's that. That's the one that and the way you were saying to link it, if there could be just a little explanation. Yeah, in the sentence you detect the data (P4)<br><br>I'm not clear if it logs it automatically…or if you have to do it manually (P2)<br><br>is this just showing me my snack time? Or is this all my exercise time? I don't know if there's a differentiation… so clarifying that would be |

| | | |
|---|---|---|
| | | interesting (P3) |
| | Make it easier to manually add activities | Because manually changing it like that. I'm not gonna do it… but your regular person who can't even be bothered to walk briskly up to the shops is not going to bother to do that, right? Why would they? (P5) |
| Recognition and rewards | Recognise longer bouts | But it's just measuring the snacks because I might think, well, I did a three hour walk and it's not registering. Maybe the app is broken or it's not showing me that that's so clarifying that would be interesting (P3) |
| | | If it is, it's not eligible, so it needs to come up saying not doesn't meet criteria or this is not snack...Because I could make myself feel a lot better by adding them all (P5) |
| Customization | | And so if your system when you put your age in can give you what a normal, healthy person of that age does, that's a bit fairer, isn't it? Cause why should an 80-year-old compete with a 70-year-old (P5) |

*Note*. P refers to participant ID
