## Supplementary material for "Promoting Vigorous Intermittent Lifestyle Physical Activity (VILPA) in middle-aged adults: An evaluation of the MovSnax mobile app": Table S5

Table S5. Part C Socio-demographic and health characteristics of participants (*N*=35)

| | Values n (%) |
|---|---|
| **Sex** | |
| Female | 28 (80) |
| Male | 7 (20) |
| **Age (years)** | |
| 40-45 | 5 (14.29) |
| 46-50 | 4 (11.43) |
| 51-55 | 6 (17.14) |
| 56-60 | 9 (25.71) |
| 60-65 | 11 (31.43) |
| **Ethnicity** | |
| Oceanian* | 19 (54.3) |
| Southeast Asian | 1 (2.9) |
| Northeast Asian | 1 (2.9) |
| Southern and Central Asian | 1 (2.9) |
| Northwest European | 6 (17.1) |
| Southern and Eastern European | 2 (5.7) |
| Other | 5 (14.3) |
| **Subjective socio-economic status** | |
| 1 (lowest) | 1 (2.90) |
| 2 | 0 (0) |
| 3 | 2 (5.90) |
| 4 | 1 (2.90) |
| 5 | 3 (8.60) |
| 6 | 7 (20) |
| 7 | 9 (25.70) |
| 8 | 4 (11.4) |
| 9 | 1 (2.90) |
| 10 (highest) | 2 (5.70) |
| **Highest level of education** | |
| Year 11 or below (incl. certificate I/II) | 2 (5.70) |
| Diploma/Advanced Diploma/Vocational training | 7 (20) |
| Bachelor/Masters/Graduate Diploma | 22 (62.90) |
| Doctoral degree/PhD | 2 (5.70) |
| Other | 2 (5.70) |
| **Marital status** | |
| Never married | 3 (8.60) |
| Married/de facto | 27 (77.10) |
| Separated/divorced | 5 (14.30) |
| **Work status** | |
| Full-time | 12 (34.30) |
| Part-time | 9 (25.70) |
| Retired | 8 (22.90) |
| Other | 6 (17.10) |
| **Current health status** | |
| Poor | 1 (2.90) |

| | |
|---|---|
| Fair | 4 (11.40) |
| Good | 11 (31.40) |
| Very good | 17 (48.60) |
| Excellent | 2 (5.70) |
| **Health issues** | |
| Yes | 18 (51.40) |
| No | 17 (48.60) |
| **Body Mass Index** | |
| <25 | 13 (37.14) |
| >25-<30 | 10 (28.57) |
| >30 | 10 (28.57) |
| **Weight perception** | |
| Very underweight | 0 (0) |
| Slightly underweight | 1 (2.90) |
| About the right weight | 10 (28.60) |
| Slightly overweight | 11 (31.40) |
| Very overweight | 13 (37.10) |

*n=18 identified as Australian; n=1 as Aboriginal/Australian South Sea Islander
