## Supplementary material for "Promoting Vigorous Intermittent Lifestyle Physical Activity (VILPA) in middle-aged adults: An evaluation of the MovSnax mobile app": Table S6

Table S6. Part D coding framework and findings

| Codes | Sub-codes | Example quotes |
|---|---|---|
| App concept | Core concept<br>Focus on short achievable activities<br>Uniqueness of concept (promoting quick and efficient ways to improve health)<br>Acknowledgements of small efforts | …once the tweaks have been made to it, it's a good concept (P2)<br><br>I don't care about steps. But this movement it made me realize that the speed of the steps is Yeah, interesting info. It is valuable. It's beneficial, I think. (P9)<br><br>I like that it was acknowledging any effort you put into…physical activity. And then it could be added as a report. Moving so it didn't have to be a gym workout (P13)<br><br>I love the idea. I mean, it's a great idea and if I could get it to work, I probably would use it. (P15) |
| User experience | Layout and design<br>User interface<br>Functionality<br>Required effort<br>Transparency<br>Clarity and knowledge<br>Navigation | I like how it's fairly simple. And pretty clean interface, so it's not super complex like an example of an app that I've used because I needed to use it, the lose it app. (P4)<br><br>Nice what was on the screen seemed pleasant and user-friendly. (P13)<br><br>It was just really easy to use. Like visually just easy to find your way around. (P7)<br><br>Yeah, I mean even graphically it wasn't entertaining. It you know it doesn't have any Yeah, it doesn't it's not user friendly…It doesn't make it easy to look at. (P8)<br><br>And the notifications that it sends out are pretty. Easy to read and easy to understand. (P1)<br><br>I thought it was really good. I actually went back to use it to get that come from the app. So. But yeah it was really motivating at the time. (P7)<br><br>The intuitiveness of it. And then people being able to look back. They haven't had to do anything they can see, I think is a good thing. (P5)<br><br>I think the user interface could do with…larger icons, larger menu items. (P2) |

Definitely work on the interface to make it, more user-friendly. (P2)

Bring in a reset password option. (P1)

I felt that maybe there was if there was some kind of explanation. As to on the same screen. Not somewhere else but on the same screen to sort of elaborate what needs to be done. I thought that would be helpful. So that's what I thought. (P12)

It seemed I was getting too much data to have to deal with if that makes sense for where I was at relative to where I was at. (P4)

I didn't like it. I. It didn't feel intuitive and I didn't feel like I was really getting any value. (P6)

I didn't understand how to use it at all. So I didn't basically very much. (P8)

So yeah, it's very limiting way of recording what you do. Without…any way to expand io tot if there is a way to expand it because I actually did try like, you know, click through screens and try two different things and I couldn't find the way if there is a way to do it…it's really very difficult (P6)

So it seemed like the effort of recording those short snaps of activity…Is just a very big effort for very little gain. And that's why I thought that, you know, it just gives so little for the effort that that's why I scored it minimal. (P6)

It takes a bit of effort and I felt that adding data manually adding data and changing the detected data was, hard. (P12)

But there was nothing to explain that and it first of all when I started using it I thought, well what action am I meant to do you know, it doesn't tell me and I was going through, you know, trying to find the answer to that question on the app and it wasn't there…So I think it would have been useful to have a little bit more preliminary information just explaining…those sorts of things (P14)

| | | |
|---|---|---|
| | | I just wanted to tell me what I've done. And like where I need to be going, what direction I need to be heading in with my vigorous movement next…You know, some sort of running total or something. I don't want to be adding things if that's what I'm supposed to be doing (P15)<br><br>Oh, I really didn't understand it at all. And I found it very not user-friendly (P8)<br><br>…this one, sop where, had the option of pressing a button saying manually add data? How is that designed to work? Cause I couldn't work that out. You couldn't do it retrospectively (P9)<br><br>I didn't know what I was supposed to be doing, what it was telling me (P15) |
| Customization | Options (e.g., font size)<br>User input<br>Person-app fit<br>Thresholds for activity detection<br>Activity categorization (e.g., what counts as a movement snack)<br>Tailoring to individual preferences | I couldn't see why I could customize a great deal. I think I made the font smaller because I literally because I have my phone settings to No, bold, and large type. (P9)<br><br>I think I probably do increase my movement just because I'm a fairly competitive person and it's always that effective (P3)<br><br>Sort of information about how I was going compared to other people in my age, like it didn't sort of say, you know, this is what most people are doing or you're behind the average for 60-year-old women kind of thing or you're doing better than most 60-year-old women or compared to other people that are using the. So there's no comparison with anybody else. Or with any sort of standard group or benchmark. Which I probably would have liked. (P10)<br><br>So again, if you're looking at that age bracket, just consider the size of the text (P13) |
| User engagement | Potential to impact PA<br>Adding an extra challenge to exercise routine | But it did keep me, entertained because I was interested to see at the end of the day. (P9) |

| | | |
|---|---|---|
| | Fun and stimulating<br>Setting challenges<br>Motivational messages<br>Monthly reports<br>Visual presentation of activity levels<br>Awareness of including movement throughout the day<br>Encouragement and feedback<br>Prompts and reminders<br>Interactive features<br>Rewards<br>Notifications<br>Reinforcement | It was easy to use. That was good. I, it did motivate me to work harder. And I did the 5 nearly every day…I did the 5 high intensity moves. So, and I wouldn't have done that without using the app. (P10)<br><br>I was more motivated to try and achieve. That achieve some data being detected and to achieve some activities. Okay. Yes. So it did increase the motivation. (P13)<br><br>But yeah, that type of thing so that you know what you're trying to do and it's really easy to do it and you can see how you're progressing (P4)<br>So there was kind of nothing apart from the reporting the numbers that…kept you engaged I guess (P3)<br><br>It didn't really feel like it was getting me to develop a more happy pattern of incidental exercised. And then now I think about it, there will also no like, G or well done messages or you know little silver and gold medals….So there is no kind of inherent reward or a sense of achievement (P3)<br><br>It would be awesome if it reminded you to just quickly go. Do a quick burst of activity (P1)<br><br>And also, there was a better way to quantify, well, what does all this mean that I've done these movement snacks. So what exactly have I achieved? (P4)<br><br>I mean it would be good if it was linked to you know reward points or something like that that you could cash in for (P11)<br><br>It didn't, if you reached a certain like your goal. Suppose you had your 5, you said you go for 5, high intensity things today. When I got 5, it didn't, you know, have fireworks or anything exciting. It just, yeah, you've got your goal kind of thing (P10)<br><br>So that was my main frustration with the app. That you're not getting positive feedback, you're not getting that sort of rewarding thing of, hey, well done, you know, you've ticked your boxes for today (P14) |

| | | |
|---|---|---|
| | | And make it a bit more response, you know, give you a bit more some cheers if you do well or. Yeah. Remind you to get up and do something if you're not doing anything (P10)<br><br>My assumption was that the app would send notifications with suggestions of exercise snacks (P2)<br><br>Like I would like it to have more ability to prompt you to do things and to have notifications and it has started that first survey the app has, started giving me notifications" (P1) |
| Transparent and meaningful tracking | Recording bursts of activity<br>Picking up various movements<br>Tracking daily activity<br>Adding activities<br>Tracking specific exercises<br>Tracking additional activities<br>Adding other types of activities<br>Adding features to record weight or other health metrics<br>Detection of activity | Yeah, it's a For me once you start getting a lot of numbers like I think you know 39 or 42 or even 30 movement snacks in a day…is too many to really. Be meaningful. (P3)<br><br>I like the fact that it was quite sensitive and it was picking up a lot of data! (P12)<br>So it was interesting to see how that shot up as vigorous movement because I don't consider it vigorous. I just think I'm going to one class to the next. But obviously, if I think about it, you know, now that I'm thinking about it, it makes sense (P9)<br><br>It just wasn't being counted and I don't understand why it wasn't whether I maybe as you said it has to be a certain speed or something like that…I'm not sure but I just found it a bit frustrating (P14)<br><br>It seemed I was getting too much data to have to deal with if that makes sense for where I was at relative to where I was at (P4)<br><br>I didn't have any trouble using it, I don't think. So it seemed to all hook up connect and it was recording everything correctly (P10) |

*Note*. P refers to participant ID
