## Supplementary material for "Promoting Vigorous Intermittent Lifestyle Physical Activity (VILPA) in middle-aged adults: An evaluation of the MovSnax mobile app": Declarations

Ethics approval and consent to participate: Ethics approval was obtained by the University of Sydney’s Human Research Ethics Committee (ethics approval no: 2021/533). Participants were informed about the study and had the opportunity to ask questions prior to agreeing to take part. All participants signed written consent forms prior to participating in the study.

Consent for publication: Not applicable

Availability of data and materials: The datasets used and analysed during the present study are available from the corresponding author upon reasonable request.

Competing interests: The authors report that they have no competing interests.

Funding: Australian National Health and Medical Research Council (APP1180812). The funder had no role in the conduct of the study.

Authors’ contributions: Conceptualization: CT-N, AG, AH, JPG, CT-L, AK, NJ, CM, JYC, ES; Methodology: CTN, AG, JYC, ES; Formal analysis: CT-N, AG, JYC; Writing – original draft preparation: CT-N; Writing – review and editing: CT-N, AG, AH, JPG, SS, CT-L, AK, NJ, CM, JYC, ES.

Acknowledgements: We would like to acknowledge the financial support of the Australian National Health and Medical Research Council (APP1180812), and the participants who took part in the study.

Electronic supplementary material:

Additional File 1: Table S1. Part A coding framework and findings (pdf)

Additional File 2: Table S2. Part B coding framework and findings (pdf)

Additional File 3: Table S3. Part C socio-demographic and health characteristics of participants (pdf)

Additional File 4: Table S4. Part D coding framework and findings (pdf)

Additional File 5: Table S5. Part C Socio-demographic and health characteristics of participants (*N*=35)

Additional File 6: Table S6. Part D coding framework and findings
